# HIV Treatment and Program Preferences Among Ryan White Clients in New York City in the Era of Long-Acting Injectable ART: A Discrete Choice Experiment

**DOI:** 10.64898/2026.02.13.26346257

**Authors:** Rebecca Zimba, Elizabeth A. Kelvin, Sarah Kulkarni, Jennifer Carmona, Tigran Avoundjian, Connor Emmert, Meghan Peterson, Mary Irvine, Denis Nash

**Author notes:** **Corresponding author**: Rebecca Zimba.

## Abstract

**Introduction:** Despite improvements in treatment for people living with HIV (PWH), adherence remains a challenge for many. In this study we aimed to identify preferences for daily pill or long-acting injectable (LAI) antiretroviral therapy (ART) and for possible treatment package features, among PWH enrolled in Ryan White HIV/AIDS Program Part A (RWPA) Medical Case Management (MCM) programs.

**Methods:** Participants were recruited from six MCM programs from across the New York RWPA eligible metropolitan area (the five boroughs of New York City and Rockland, Putman, and Westchester counties). We developed a discrete choice experiment (DCE) with four attributes: (1) Type of ART Medication (daily pills or LAI), (2) Service Location and Mode, (3) Support, and (4) Rewards. We used an alternative-specific design in which the levels for the last three attributes were dependent on levels within the first (Type of ART). Latent class multinomial logit analysis (LCA) was used for preference estimation and hypothesis-free investigation of preference heterogeneity.

**Results:** From June 2022 through January 2023, 200 New York RWPA MCM clients completed the DCE. We selected a two-group LCA solution. A majority of participants had a higher preference for LAI regimens compared to daily pills (n=114 [57%] versus n=86 [43%]). Those who preferred LAI ART were younger (median age 49 versus 58.5 years, p<0.001), less likely to identify as straight/heterosexual (69% versus 82%, p=0.03), and more likely to identify as Latino/a (54% versus 30%; p<0.001). Preferences for service locations/mode, supports, and rewards were similar across LCA groups. Participants who preferred LAI ART were more likely to have heard of LAI ART before the survey (59% versus 41%, p=0.012). Overall, only 4% of participants self-reported having tried LAI ART.

**Conclusions:** Assessing preferences among groups under-represented in clinical trials is essential to effective and equitable real-world implementation of innovative treatment options. Our study found that there were distinct groups that differed in their preferred ART regimen type and that New York RWPA MCM clients had limited familiarity with LAI ART. To inform regimen selection, we began pilot-testing educational materials and a patient-provider decision-making tool in 2023.

## Introduction

Treatment for people living with HIV (PWH) has improved over time and the current standard involves a single, well-tolerated, daily pill. However, adherence remains a challenge for many and is an obstacle to realizing the Ending the HIV Epidemic goal to reduce new HIV infections by 90% by 2031.^1,2^ In 2022, among all persons diagnosed with HIV in 57 United States (US) jurisdictions and all US states with complete viral load (VL) reporting, only 65.1% had evidence of viral suppression (VS; VL <200 copies/mL) on their most recent test result; the proportion was lower for Black PWH (60.5%) than for Latino/a (64.3%) or White PWH (70.8%).^3^ Racism, poverty, stigma, trauma, and incarceration can all contribute to treatment barriers like unstable housing, mental illness, and substance use disorders.^4–7^

In January 2021, the US Federal Drug Administration (FDA) approved Cabenuva (cabotegravir/rilpivirine), as a stand-alone long-acting injectable (LAI) monthly antiretroviral treatment (ART) regimen;^8^ in February 2022, a bimonthly injectable Cabenuva regimen was approved.^9^ Cabenuva is approved for adolescent and adult PWH who are virally suppressed and ≥35kg. The monthly or bimonthly dosing of LAI ART may circumvent some of the common barriers to adherence, such as housing instability, stigma, mental health, and substance use disorders, by decoupling VS from daily medication-taking. In so doing, LAI ART could narrow racial/ethnic and socioeconomic disparities in VS and mortality. However, uptake of Cabenuva has been slow.^10,11^

Qualitative studies have found that some patients like the convenience of LAI ART and its potential to prevent accidental HIV status disclosures, but others have expressed concerns over effectiveness and injection side effects, and have recognized that many of the same barriers that interfere with adherence to daily pill ART may also impact keeping injection appointments.^12–15^ PWH acceptability of or openness to LAI ART ranged from 40%–74% in studies conducted before FDA approval.^16–20^ In a study of uptake in routine care September–December 2021, 31.4% of PWH elected to initiate LAI ART.^21^

Biomedical innovations historically have most benefitted those who are already relatively advantaged, and can even exacerbate health disparities.^22–24^ The slow uptake of pre-exposure prophylaxis (PrEP)^25^ and lower awareness of, use of, and adherence to PrEP among women and people of color^26–30^ highlights the need to integrate user perspectives into the rollout of new biomedical products to promote equitable impact. For LAI ART, it is important to understand perceptions and preferences among PWH in diverse care settings and communities hardest hit by HIV, which are often underrepresented in clinical trials and may have been conditioned to view new biomedical technologies with distrust.^31–34^

In this study we used a discrete choice experiment (DCE) to identify preferences for LAI or daily pill ART and for possible treatment package features among PWH enrolled in New York Ryan White HIV/AIDS Program Part A (RWPA) Medical Case Management (MCM) programs, which are designed to address barriers to adherence.

## Methods

### Study setting & sample

Participants were recruited from six New York RWPA MCM programs, representing different geographic regions (three programs across the five boroughs of New York City [NYC] and three programs in Rockland, Putman, and Westchester Counties), and all organizational settings where RWPA MCM services are provided.^35^ A high proportion of clients of New York RWPA services are low-income (88.4% are ≤138% of the 2023 Federal Poverty Level) and Black (47.9%) or Latino/a (39.3%).^36,37^ The Ryan White HIV/AIDS Program is an essential resource for reducing HIV-related health disparities in the US and for scaling up evidence-based practices to optimize care continuum outcomes.^38^ Since 1990 this federal program has provided funding to cover medical and support services for PWH, serving over half of PWH in the US.^38–42^

### DCE design and pilot testing

In Fall 2021, we conducted focus groups to inform the design of the DCE.^43,44^ We held two focus groups with five clients (one via Zoom and one in-person), and three role-specific focus groups with 16 providers (all via Zoom): one with program directors, another with care coordinators and patient navigators, and a third with prescribing providers. (See Supplemental Table 1 for focus group participant demographic characteristics.) As we developed the attributes and levels for the DCE, we incorporated focus group findings about the perceived advantages and disadvantages of LAI ART, potential barriers to and facilitators of LAI ART uptake, and implementation considerations. We also considered results from previous studies, including clinical trials focused on LAI ART.^13,45^

The DCE was designed and deployed using Lighthouse Studio (version 9, Sawtooth Software, Provo, UT) with four attributes: (1) Type of ART Medication (daily pills or LAI), (2) Service Location and Mode, (3) Support, and (4) Rewards. To ensure plausible combinations of treatment-package features, we used an alternative-specific design in which the levels for the last three attributes depended on levels within the first.^46,47^ In April 2022, we pilot tested the survey with ten clients enrolled in a RWPA MCM program that would not be participating in the final client survey and gathered feedback on user experience, clarity of question wording, and ease of taking the survey on a mobile device. Pilot testers were compensated with a $25 gift card. The final DCE design contained twelve choice sets, each including two alternatives plus a None option in case neither alternative was preferred (Supplemental Table 2 and Supplemental Figure 1).

We generated random tasks using the balanced overlap design option in Lighthouse Studio, which achieves level balance and orthogonality while also allowing for some overlap within attributes across the alternatives in a choice task. Participants were asked to choose their preferred alternative for each choice set presented, implicitly making trade-offs between the desirable and undesirable characteristics of each option.

To contextualize the DCE results, we included questions covering prior experience with LAI ART, comfort taking pills, and comfort with the idea of injections for HIV treatment. We also included the Acceptability of Intervention Measure (AIM)^48^ for LAI ART, comprising four questions scored from 1 (“Completely disagree”) to 5 (“Completely agree”), which were then averaged.

### Sample Size, Recruitment, and Data Collection

We set our target sample size to 200 participants, which was sufficient to estimate main effects with precision. (See Supplemental Materials for additional details.) RWPA MCM clients were eligible to participate if they were ≥18 years of age (adults) and enrolled in RWPA MCM services at one of the six partnering agencies. Between June 2022–January 2023, program staff at partnering MCM agencies contacted eligible clients about the opportunity to participate. Those who were interested were given the URL and unique login codes for the survey, which they could complete onsite or elsewhere using any internet-connected device. Clients were required to review a consent form and provide electronic informed consent before beginning the survey. Those who completed the survey received a $25 gift card for their participation.

Survey data were merged with NYC and Tri-county HIV surveillance registry data and RWPA MCM records from the NYC Health Department’s programmatic data source (eSHARE). The study methods were reviewed and approved by the Institutional Review Board of the New York City Department of Health and Mental Hygiene (Protocol #20-096).

### Statistical Analysis

We described study participants overall and compared with other adult New York RWPA MCM clients, assessing the statistical significance of differences using chi-square tests or Fisher’s exact tests if expected cell counts were <5 for categorical variables, and two-sided t-tests or Wilcoxon rank sum tests for continuous variables. We used Lighthouse Studio’s Analysis module to conduct a latent class multinomial logit analysis (LCA) to estimate HIV treatment program preferences (zero-centered part-worth utilities^46,47^) and for hypothesis-free investigation of preference heterogeneity.^49^ We chose the LCA solution balancing improvements in model fit statistics (percent certainty [a pseudo-R^2^], Akaike’s Information Criterion, and Bayesian Information Criterion) with group sizes, interpretability, and relevance of group differences.^50^ We gauged response quality through consistency metrics (respondent speed, root likelihood statistic, and straight-lining), and conducted a sensitivity analysis excluding poor-quality responses.^51^

## Results

### Description of study participants

From June 2022–January 2023, 200 New York RWPA MCM clients completed the survey. Participants’ median age was 55 years and median time since RWPA MCM enrollment was 31.2 months. Most participants were Black or Latino/a, and most identified as men. One-quarter self-reported imperfect adherence to daily ART, and 86% were virally suppressed (Table 1).

**Table 1.** Characteristics of clients who completed the APPLI DCE compared to all other New York City and Tri-County MCM clients ≥18 years of age.

|  | Overall<br>N = 2,669<sup>†</sup> | DCE completion status |  | p-value |
| --- | --- | --- | --- | --- |
|  |  | Completed<br>DCE<sup>†,‡</sup><br>n=200 (7.5%) | All other MCM<br>clients<sup>†</sup><br>n=2469 (93%) |  |
| DEMOGRAPHIC VARIABLES |  |  |  |  |
| Age in years | 51 (37, 61) | 55 (41, 62) | 51 (37, 61) | 0.069<sup>§</sup> |
| Missing/Unknown | 2 | 2 | 0 |  |
| Age group in years |  |  |  | 0.2 |
| 18-29 | 213 (8.0%) | 17 (8.6%) | 196 (7.9%) |  |
| 30-39 | 558 (21%) | 30 (15%) | 528 (21%) |  |
| 40-49 | 487 (18%) | 33 (17%) | 454 (18%) |  |
| 50-59 | 610 (23%) | 47 (24%) | 563 (23%) |  |
| 60+ | 799 (30%) | 71 (36%) | 728 (29%) |  |
| Missing/Unknown | 2 | 2 | 0 |  |
| Self-reported gender |  |  |  | 0.017 |
| Cisgender man | 1,652 (62%) | 117 (59%) | 1,535 (62%) |  |
| Cisgender woman | 899 (34%) | 79 (40%) | 820 (33%) |  |
| Transgender, gender-non-conforming, or non-binary | 115 (4.3%) | 2 (1.0%) | 113 (4.6%) |  |
| Missing/Unknown | 3 | 2 | 1 |  |
| Sexual identity |  |  |  | <0.001 |
| Homosexual, bisexual, pansexual, or queer | 958 (37%) | 50 (25%) | 908 (38%) |  |
| Straight/heterosexual | 1,616 (63%) | 147 (75%) | 1,469 (62%) |  |
| Missing/Unknown | 95 | 3 | 92 |  |
| Race/Ethnicity<sup>††</sup> |  |  |  | 0.3<sup>‡‡</sup> |
| Latino/a | 1,158 (44%) | 87 (44%) | 1,071 (43%) |  |
| Asian/Pacific Islander | 51 (1.9%) | 0 (0%) | 51 (2.1%) |  |
| Black | 1,291 (48%) | 99 (50%) | 1,192 (48%) |  |
| White | 120 (4.5%) | 9 (4.5%) | 111 (4.5%) |  |
| Other/Multiracial | 42 (1.6%) | 3 (1.5%) | 39 (1.6%) |  |
| Missing/Unknown | 7 | 2 | 5 |  |
| Primary language |  |  |  | 0.2 |
| English | 1,647 (62%) | 116 (59%) | 1,531 (62%) |  |
| Spanish | 821 (31%) | 67 (34%) | 754 (31%) |  |
| Haitian Creole | 75 (2.8%) | 9 (4.5%) | 66 (2.7%) |  |
| Other primary language | 121 (4.5%) | 6 (3.0%) | 115 (4.7%) |  |
| Missing/Unknown | 5 | 2 | 3 |  |
| Highest level of schooling completed |  |  |  | 0.7 |
| High school/GED or lower | 1,876 (72%) | 137 (71%) | 1,739 (72%) |  |
| Greater than high school/GED | 737 (28%) | 57 (29%) | 680 (28%) |  |
| Missing/Unknown | 56 | 6 | 50 |  |
| Employment status |  |  |  | 0.010 |
| Paid Employment | 782 (29%) | 75 (38%) | 707 (29%) |  |
| No Paid Employment | 1,416 (53%) | 86 (44%) | 1,330 (54%) |  |
| Out of Workforce | 457 (17%) | 36 (18%) | 421 (17%) |  |
|  |  | Completed<br>DCE<sup>†,‡</sup><br>n=200 (7.5%) | All other MCM<br>clients<sup>†</sup><br>n=2469 (93%) |  |
| Missing/Unknown | 14 | 3 | 11 |  |
| **Housing status<sup>§§</sup>** |  |  |  | 0.10<sup>¶</sup> |
| Stably Housed | 2,156 (81%) | 169 (85%) | 1,987 (81%) |  |
| Unstably Housed or Homeless | 508 (19%) | 29 (15%) | 479 (19%) |  |
| Missing/Unknown | 5 | 2 | 3 |  |
| **MCM ENROLLMENT AND AGENCY VARIABLES** |  |  |  |  |
| **Time since MCM enrollment in months** | 27.7 (11.6, 44.3) | 31.2 (11.8, 45.8) | 27.4 (11.6, 44.2) | 0.2<sup>§</sup> |
| Missing/Unknown | 2 | 2 | 0 |  |
| **Agency category** |  |  |  | <0.001<sup>¶</sup> |
| Community health center | 1,016 (38%) | 107 (54%) | 909 (37%) |  |
| Community-based organization | 516 (19%) | 59 (30%) | 457 (19%) |  |
| Private hospital | 713 (27%) | 0 (0%) | 713 (29%) |  |
| Public hospital | 424 (16%) | 34 (17%) | 390 (16%) |  |
| **Agency location** |  |  |  | <0.001<sup>¶</sup> |
| Bronx | 475 (18%) | 37 (19%) | 438 (18%) |  |
| Brooklyn | 544 (20%) | 34 (17%) | 510 (21%) |  |
| Manhattan | 911 (34%) | 25 (13%) | 886 (36%) |  |
| Queens | 277 (10%) | 0 (0%) | 277 (11%) |  |
| Staten Island | 67 (2.5%) | 0 (0%) | 67 (2.7%) |  |
| Tri-County | 395 (15%) | 104 (52%) | 291 (12%) |  |
| **MENTAL HEALTH AND SUBSTANCE USE VARIABLES** |  |  |  |  |
| **Psychiatric diagnoses (lifetime history, not mutually exclusive)<sup>¶¶</sup>** |  |  |  |  |
| Depression or anxiety disorder | 1,127 (42%) | 96 (48%) | 1,031 (42%) | 0.067<sup>¶</sup> |
| Psychosis or bipolar disorder | 338 (13%) | 30 (15%) | 308 (12%) | 0.3<sup>¶</sup> |
| Post-traumatic stress disorder | 200 (7.5%) | 20 (10%) | 180 (7.3%) | 0.2<sup>¶</sup> |
| Other psychiatric diagnosis | 96 (3.6%) | 5 (2.5%) | 91 (3.7%) | 0.4<sup>¶</sup> |
| Missing/Unknown (for all psychiatric diagnoses) | 5 | 2 | 3 |  |
| **Recent problem substance use<sup>†††</sup>** | 480 (19%) | 18 (10%) | 462 (19%) | 0.003<sup>¶</sup> |
| Missing/Unknown | 77 | 21 | 56 |  |
| **HIV CLINICAL AND TREATMENT VARIABLES** |  |  |  |  |
| **Time since HIV diagnosis in years** | 15.7 (6.7, 25.8) | 18.2 (8.8, 27.5) | 15.5 (6.5, 25.3) | 0.004<sup>§</sup> |
| Missing/Unknown | 58 | 15 | 43 |  |
| **ART regimen proximal to date of survey** |  |  |  | 0.013<sup>‡‡</sup> |
| Daily oral ART | 2,504 (94%) | 196 (99%) | 2,308 (94%) |  |
| LAI ART | 13 (0.5%) | 0 (0%) | 13 (0.5%) |  |
| Other, unspecified ART | 74 (2.8%) | 2 (1.0%) | 72 (2.9%) |  |
| Not on ART | 67 (2.5%) | 0 (0%) | 67 (2.7%) |  |
| Missing/Unknown | 11 | 2 | 9 |  |
| **Percent adherence<sup>‡‡‡</sup>** |  |  |  | <0.001<sup>¶</sup> |
| Below 50% | 294 (12%) | 7 (3.6%) | 287 (13%) |  |
| 50-69% | 112 (4.5%) | 6 (3.1%) | 106 (4.7%) |  |
| 70-89% | 171 (6.9%) | 9 (4.6%) | 162 (7.1%) |  |
| 90-94% | 195 (7.9%) | 13 (6.6%) | 182 (8.0%) |  |
| 95-99% | 164 (6.7%) | 10 (5.1%) | 154 (6.8%) |  |

|  | DCE completion status |  |  | p-value |
| --- | --- | --- | --- | --- |
|  | Overall<br>N = 2,669<sup>†</sup> | Completed<br>DCE<sup>†,‡</sup><br>n=200 (7.5%) | All other MCM<br>clients<sup>†</sup><br>n=2469 (93%) |  |
| 100% | 1,527 (62%) | 151 (77%) | 1,376 (61%) |  |
| Missing/Unknown | 41 | 0 | 41 |  |
| **Receipt of directly observed therapy within MCM<sup>§§§</sup>** | 673 (25%) | 50 (25%) | 623 (25%) | >0.9<sup>¶¶</sup> |
| **Most recent ART regimen<sup>¶¶¶¶</sup>** |  |  |  | <0.001<sup>¶¶</sup> |
| Daily oral ART | 2,439 (92%) | 175 (88%) | 2,264 (92%) |  |
| LAI ART | 91 (3.4%) | 18 (9.1%) | 73 (3.0%) |  |
| Other, unspecified ART | 62 (2.3%) | 1 (0.5%) | 61 (2.5%) |  |
| Not on ART | 66 (2.5%) | 4 (2.0%) | 62 (2.5%) |  |
| Missing/Unknown | 11 | 2 | 9 |  |
| **Viral load (copies/mL)<sup>¶¶¶¶</sup>** |  |  |  | 0.067<sup>¶¶</sup> |
| <200 | 2,168 (82%) | 171 (86%) | 1,997 (81%) |  |
| ≥200 | 492 (18%) | 27 (14%) | 465 (19%) |  |
| Missing/Unknown | 9 | 2 | 7 |  |
| **CD4 category (cells/mm3)<sup>¶¶¶¶</sup>** |  |  |  | <0.001<sup>¶¶</sup> |
| <200 | 563 (21%) | 73 (37%) | 490 (20%) |  |
| 200-349 | 383 (14%) | 23 (12%) | 360 (15%) |  |
| 350-499 | 466 (18%) | 22 (11%) | 444 (18%) |  |
| 500+ | 1,239 (47%) | 80 (40%) | 1,159 (47%) |  |
| Missing/Unknown | 18 | 2 | 16 |  |
| **Resistance testing history** |  |  |  | <0.001<sup>¶¶</sup> |
| Resistance testing in past year | 351 (13%) | 16 (8.1%) | 335 (14%) |  |
| Resistance testing >1 year ago | 1,356 (51%) | 58 (29%) | 1,298 (53%) |  |
| No resistance testing reported | 960 (36%) | 124 (63%) | 836 (34%) |  |
| Missing/Unknown | 2 | 2 | 0 |  |
| **Time since last HIV ART resistance test in months<sup>¶¶¶¶</sup>** | 42.2 (15.1, 80.9) | 54.3 (15.2, 120.8) | 42.0 (15.1, 80.3) | 0.2<sup>§</sup> |

Abbreviation: ART = antiretroviral therapy; DCE = discrete choice experiment; GED = general equivalency diploma; MCM = medical case management.
<sup>†</sup> Median (Q1, Q3); n (%)
<sup>‡</sup> DCE and programmatic records could not be linked for two clients who completed the DCE.
<sup>§</sup> Wilcoxon rank sum test
<sup>¶</sup> Pearson's Chi-squared test
<sup>¶¶</sup> Black, White, Asian/Pacific Islander, and Other/Multiracial race categories exclude Latino/a ethnicity. People with the ethnicity Latino/a are grouped in the Latino/a category regardless of their race classification
<sup>¶¶¶</sup> Fisher's exact test
<sup>§§</sup> 'Stable' defined as any permanent housing situation. 'Unstable' defined as any temporary/transitional housing situation or being homeless. Reported as of client's latest available eSHARE assessment before date of survey completion OR January 31, 2023.
<sup>¶¶¶</sup> Any report of a psychiatric diagnosis on any eSHARE assessment before the date of survey completion OR January 31, 2023.
<sup>¶¶¶¶</sup> Recent report of binge drinking, heroin, meth, cocaine, or recreational prescription drug use in last 3 months. Reported as of client's latest available eSHARE assessment before date of survey completion OR January 31, 2023.
<sup>¶¶¶¶</sup> Self-reported percent of the past 30 days on which client took their ART pills, among clients taking Daily oral ART, as of client's latest available eSHARE assessment before date of survey completion OR January 31, 2023.
<sup>§§§</sup> Includes directly observed therapy received from MCM agencies in the last decade.
<sup>¶¶¶¶</sup> As of October 28, 2025.
<sup>¶¶¶¶</sup> Most recent lab values or self-reported values recorded in eSHARE before date of survey completion OR January 31, 2023.

### Comparison of study participants with non-participating New York RWPA MCM clients

Study participants were significantly more likely to self-identify as women, to identify as straight/heterosexual, and to be employed, compared to 2,469 non-participating adult New York RWPA MCM clients across all sites (Table 1 and Supplemental Figure 2). Compared to non-participants, participants were more likely to be enrolled in an MCM program at a community-based organization or community health center; their program was more likely to be based in the Tri-County area and less likely to be based in Manhattan; and they were less likely to have self-reported recent problem substance use. Participants had been living with HIV longer, were more likely to report 100% ART adherence, to have lower numbers of CD4 cells/mm^3^, and to have no resistance testing history. (See Supplemental Table 3 for a comparison of study participants and non-participating adult New York RWPA MCM clients from partnering sites.)

### Comparison of participant characteristics by their ART preference

We selected a two-group LCA solution (see Supplemental Table 4 for model fit statistics). One group of participants had higher preferences for the daily pill ART regimen (Group 1 [n=66, 43%]); the other group preferred the LAI regimens (Group 2 [n=114, 57%]) (Table 2). Those preferring the LAI regimens, in Group 2, were younger, diagnosed more recently, and less likely to identify as straight/heterosexual, less likely to identify as Black, and more likely to identify as Latino/a than Group 1 participants. Group 2 participants were more likely report their primary language was Spanish, and more likely to report that someone helped them take the survey by reading the questions to them. The MCM programs of participants in Group 2 were more likely to be community health centers and less likely to be community-based organizations, more likely to be based in Manhattan and less likely to be based in the Bronx.

**Table 2.**
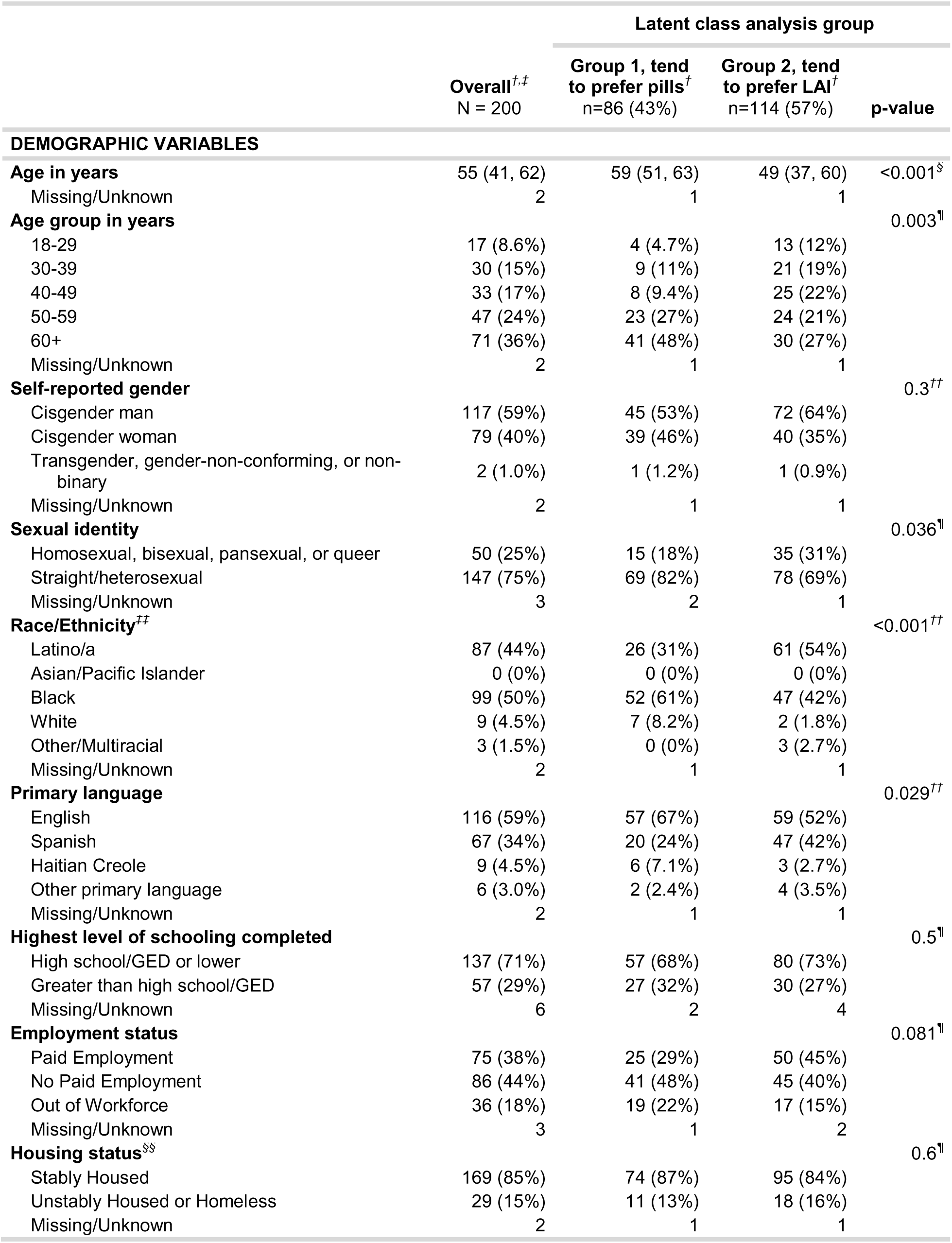
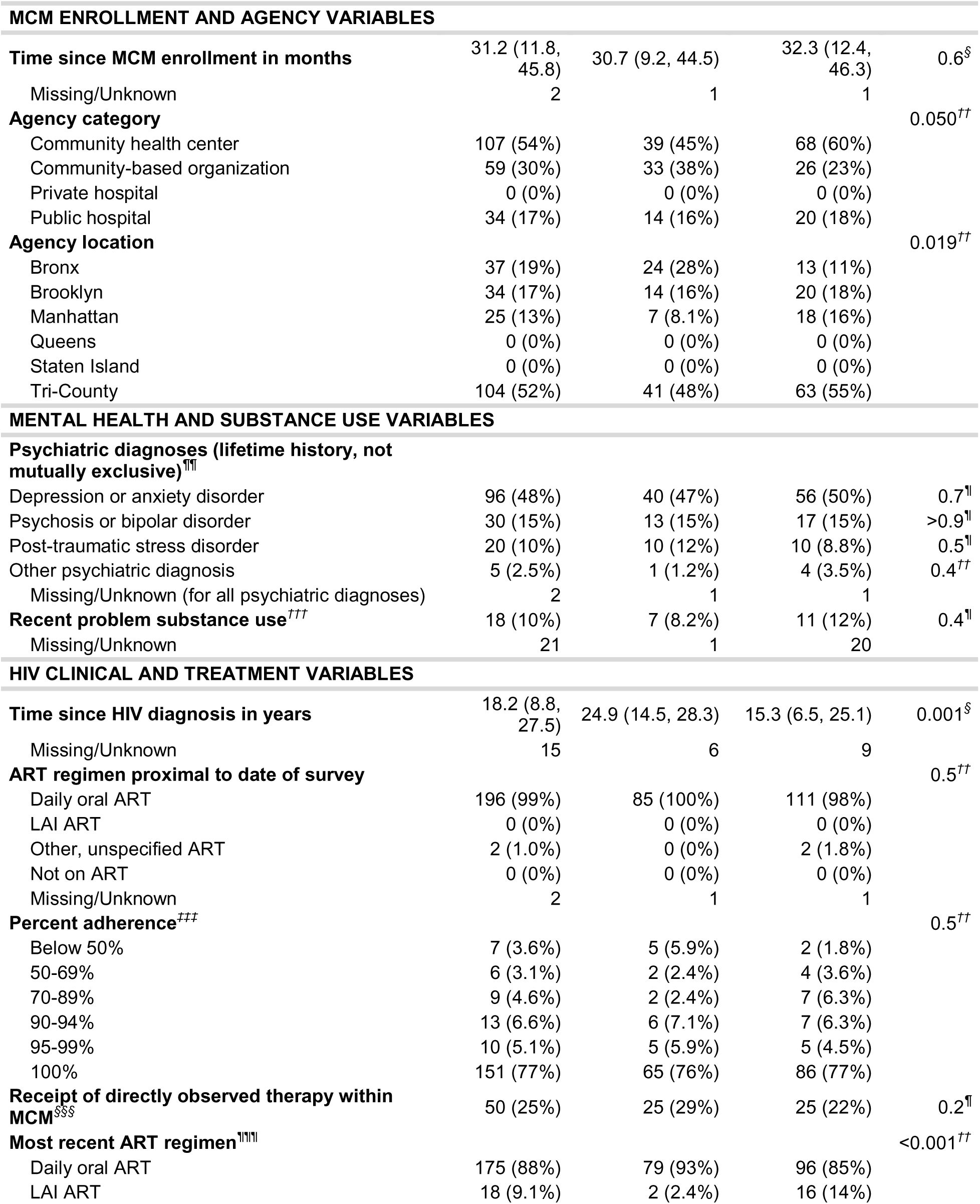
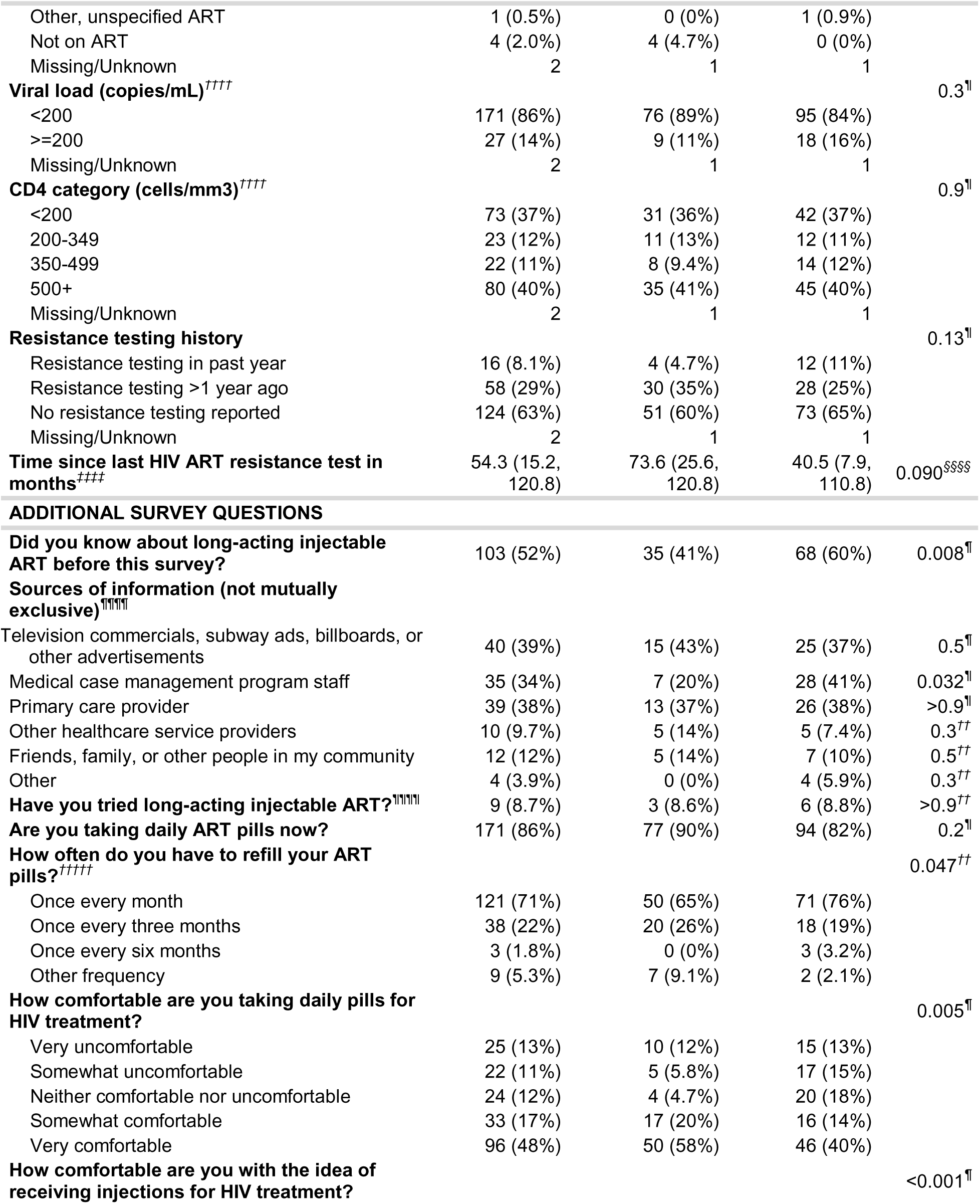
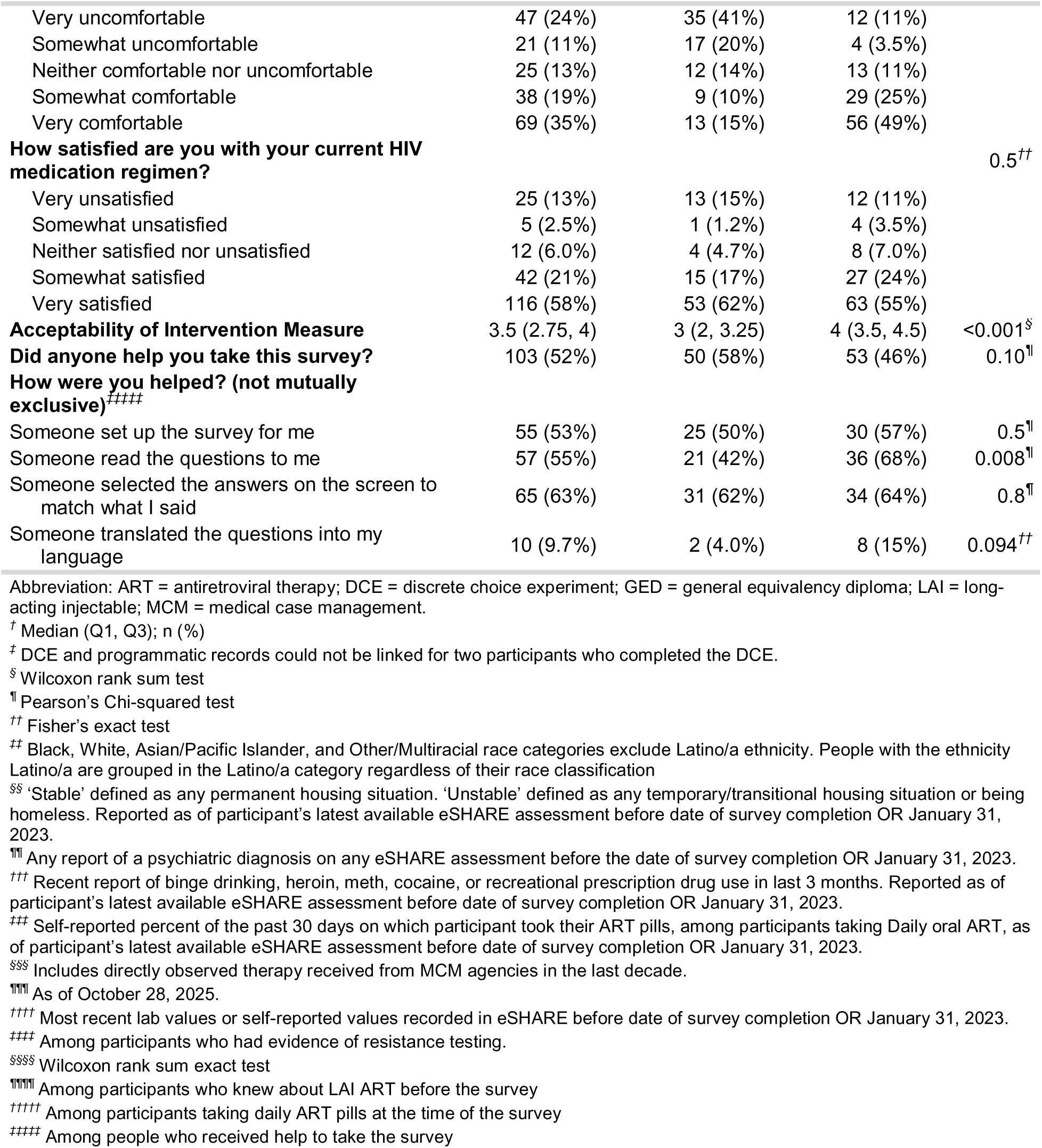
Characteristics, ART experience, knowledge & comfort among clients who completed the APPLI DCE overall and by latent class group.

Overall, half of study participants reported having heard about LAI ART before the survey, but only 4% said they had tried it (9% of those who had heard of LAI ART before). Group 2 participants were more likely to have heard of LAI ART before the survey. The most common sources of information about LAI were advertisements, MCM program staff and primary care providers. Group 2 participants were more likely to have heard about LAI ART from MCM program staff than Group 1 participants.

When asked “How comfortable are you taking daily pills for HIV treatment?” 65% of participants reported that they were somewhat or very comfortable; in contrast, when asked “How comfortable are you with the idea of receiving injections for HIV treatment?” 54% said that they were somewhat or very comfortable. Group 2 participants were less likely to report being somewhat or very comfortable with daily pill ART, and more likely to report being somewhat or very comfortable with the idea of injections for ART. Similarly, Group 2 participants had higher acceptability of LAI on the AIM items than Group 1 participants.

### DCE results

Overall, participants preferred daily pill ART over bimonthly or monthly LAI ART, and the None option had a large negative utility, indicating that most users were able to pick a preferred option from the two program alternatives (Table 3 and Figure 1). However, the two-group LCA identified groups with differing preferences: Group 1 participants preferred daily pill ART and Group 2 participants preferred bimonthly LAI ART and monthly LAI ART. Group 1 had a positive utility for the None option, and Group 2 had a large negative utility for the None option.

**Figure 1.**
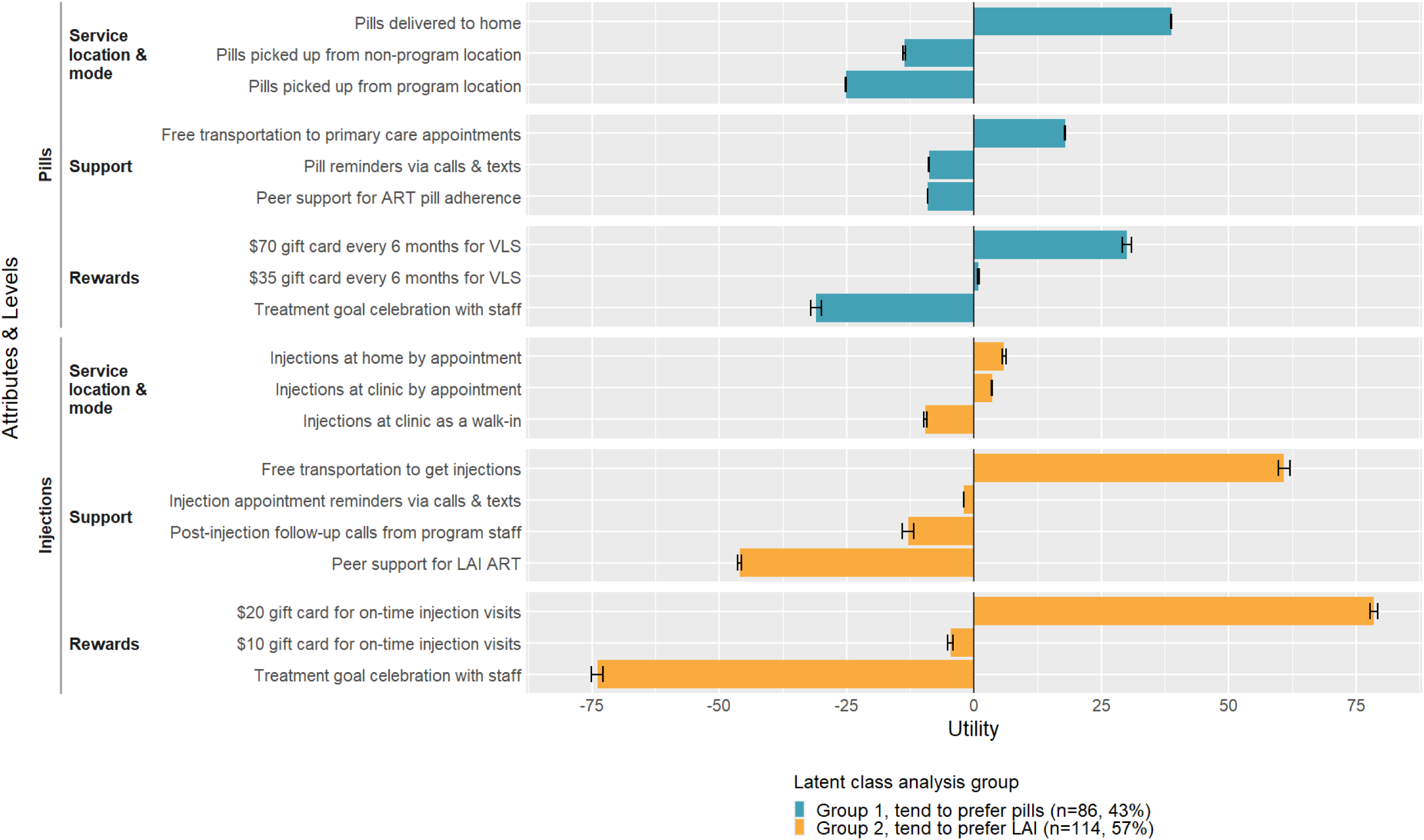
Preferences of clients from the APPLI DCE estimated with alternative-specific utilities from latent class multinomial logit regression analysis, two-group solution

**Table 3.** Preferences of clients from the APPLI DCE estimated with alternative-specific utilities from latent class multinomial logit regression analysis, two-group solution, overall and by latent class group.

| Attribute | Levels | Latent class analysis groups<sup>1</sup> |  |  |
| --- | --- | --- | --- | --- |
|  |  | Overall<br>N = 200<br>Mean (95% CI) | Group 1, tend to prefer<br>pills<br>n = 86 (43%)<br>Mean (95% CI) | Group 2, tend to prefer<br>LAI<br>n = 114 (57%)<br>Mean (95% CI) |
| Type of ART Medication | Daily oral ART pill(s) | 44.8 (23.4, 66.2) | 218.0 (215.9, 220.2) | -85.8 (-92.7, -79.0) |
|  | An ART injection in each buttock once every two months | -0.1 (-10.7, 10.5) | -86.2 (-87.5, -85.0) | 64.8 (61.8, 67.9) |
|  | An ART injection in each buttock once a month | -44.7 (-55.5, -33.9) | -131.8 (-132.8, -130.9) | 21.0 (17.2, 24.8) |
| Service Location and Mode – Pills | ART pills delivered to home | 30.7 (29.6, 31.8) | 38.8 (38.7, 38.9) | 24.6 (23.8, 25.4) |
|  | ART pills pickup from program location | -12.3 (-13.9, -10.8) | -25.1 (-25.2, -25.0) | -2.7 (-3.3, -2.0) |
|  | ART pills pickup at pharmacy or other non-program location | -18.4 (-19.0, -17.8) | -13.7 (-13.8, -13.5) | -21.9 (-22.2, -21.7) |
| Support – Pills | Free transportation provided to primary care appointments | 17.1 (16.8, 17.3) | 17.9 (17.8, 18.0) | 16.4 (16.1, 16.8) |
|  | Peer support group to help with ART pill adherence | -7.6 (-7.8, -7.4) | -9.1 (-9.1, -9.1) | -6.5 (-6.7, -6.3) |
|  | Calls and texts to remind about taking ART pills | -9.4 (-9.6, -9.3) | -8.8 (-8.9, -8.8) | -9.9 (-10.1, -9.8) |
| Rewards – Pills | \$70 gift card every 6 months for having undetectable or suppressed viral load | 62.8 (58.8, 66.8) | 30.1 (29.2, 31.0) | 87.5 (86.8, 88.1) |
| | \$35 gift card every 6 months for having undetectable or suppressed viral load | 9.9 (8.8, 11.0) | 0.9 (0.7, 1.1) | 16.7 (16.5, 16.9) |
|  | Rewards ceremony with program staff to celebrate meeting treatment goals | -72.7 (-77.8, -67.6) | -31.0 (-32.0, -29.9) | -104.2 (-105.0, -103.4) |
| Service Location and Mode – Injections | Injections by appointment at clinic | 4.5 (4.4, 4.7) | 5.8 (5.8, 5.8) | 3.6 (3.4, 3.7) |
|  | Walk-in for injections at clinic with no appointment required | -2.0 (-3.2, -0.8) | 7.9 (7.8, 8.1) | -9.5 (-9.8, -9.2) |
|  | Injections by appointment at home | -2.5 (-3.9, -1.1) | -13.8 (-13.9, -13.6) | 5.9 (5.5, 6.4) |
| Support – Injections | Free transportation provided to get injections | 28.3 (23.1, 33.6) | -14.8 (-15.6, -14.0) | 60.9 (59.8, 62.0) |
|  | Post-injection follow-up calls from program staff | 8.6 (5.1, 12.2) | 37.2 (36.9, 37.6) | -12.9 (-14.1, -11.8) |
|  | Calls and texts to remind about injection appointments | -0.9 (-1.1, -0.8) | 0.4 (0.4, 0.5) | -2.0 (-2.0, -1.9) |
|  | Peer support group for clients on or considering long-acting injectable ART | -36.0 (-37.7, -34.4) | -22.9 (-23.3, -22.4) | -46.0 (-46.4, -45.6) |
| Rewards – Injections | \$20 gift card for every injection visit in the right time frame | 65.9 (63.8, 68.0) | 49.2 (48.5, 49.9) | 78.5 (77.7, 79.3) |
| | \$10 gift card for every injection visit in the right time frame | 5.1 (3.5, 6.6) | 17.8 (17.7, 18.0) | -4.6 (-5.1, -4.0) |
|  | Rewards ceremony with program staff to celebrate meeting treatment goals | -71.0 (-71.8, -70.1) | -67.0 (-67.6, -66.5) | -74.0 (-75.1, -72.8) |
|  | None | -243.1 (-281.7, -204.4) | 72.5 (66.4, 78.6) | -481.1 (-489.0, -473.2) |

<sup>1</sup> All group comparisons statistically significant at p<0.001 using the Wilcoxon rank sum test
Abbreviations: ART = antiretroviral therapy, CI = confidence interval, LAI = long-acting antiretroviral therapy

Given the daily pill ART option, within the Service Location and Mode attribute, Group 1 participants preferred ART pills delivered to their homes over picking their pills up from a program location or from a pharmacy or other non-program location. Preferences for Support in Group 1 were highest for free transportation to primary care appointments, followed by peer support groups to help with adherence and reminder calls and texts about taking ART pills. Within the Rewards attribute, Group 1 participants preferred the $70 incentive every 6 months for having suppressed VL over the $35 incentive or the treatment goal celebration with staff.

Given the LAI ART option, within the Service Location and Mode attribute, Group 2 participants preferred injections by appointment at their homes over injections by appointment at their clinic or walk-in clinic injection visits. Preferences for Support in Group 2 were highest for free transportation to injection appointments, followed by reminder calls and texts about injection appointments, post-injection follow-up calls from program staff, and peer support groups for clients on or considering LAI ART. Within the Rewards attribute, Group 2 participants preferred the $20 incentive for every injection visit in the right time frame over the $10 incentive or the treatment goal celebration with staff.

### Sensitivity analysis

The preferences for daily pill ART in Group 1 and LAI ART in Group 2 persisted in the sensitivity analysis. The ordering of levels changed in two cases: (1) peer support became the most preferred level within the Support attribute in Group 1, switching places with reminder calls and texts about injection appointments, and (2) injections by appointment at a clinic became the most preferred level within the Support attribute in Group 2, switching places with injections by appointment at home. All other levels retained the same order of preference as in the primary analysis, though there were some differences in magnitude. (See Supplemental Table 5 and Supplemental Figure 3 for additional sensitivity analyses results.)

## Discussion

We found an overall preference for daily pill ART over LAI ART, with preference heterogeneity evident in a two-class LCA solution. Participants in Group 1 preferred daily pill ART, and participants in Group 2 preferred LAI ART, either bimonthly or monthly. This is consistent with preference variability for ART mode reported elsewhere.^16–20,52–54^ Participants preferring LAI ART tended to be younger and more likely to identify as Latino/a, as a sexual minority, and as a cisgender man, although the gender difference was not statistically significant. There was no difference in self-reported daily pill ART adherence or VS by LCA group, similar to other findings.^17^

Group 2 participants were younger and more likely to have reported recent problem substance use than Group 1 participants, though the substance use difference was not statistically significant. In some reports, PWH <30 years or in the injection drug use transmission category have been found to experience lower levels of VS on daily pill ART regimens compared to older age groups or other transmission groups.^55,56^ Our findings suggest a readiness for LAI ART in these priority populations for ending the HIV epidemic in the US, underscoring the potential of LAI ART to reduce previously persistent HIV outcome disparities and the importance of facilitating those populations’ access to LAI ART options.

Other preferences for support services were similar across LCA groups. Those in Group 1 preferred home-delivered pills and those in Group 2 would prefer home-delivered injections. Both groups preferred free rides to appointments and gift cards at the highest monetary value in recognition of demonstrated regimen adherence. Clients may experience challenges around transportation to appointments.^13,15^ which could explain their preference for home-based appointments and free transportation to appointments. Monetary incentives have become increasingly common as a means to encourage attending appointments,^13^ and to promote blood draws and achievement of VS.^57,58^ In the current study, the expressed preference for higher incentives over a celebration of VS achievement may reflect the degree of financial hardship among RWPA MCM clients.

Overall, participants reported limited familiarity with LAI ART; those in Group 2 were more likely to have heard about LAI ART before the survey. Experience with other healthcare services (e.g., hormonal contraceptives, HIV self-testing) has demonstrated that presenting people with treatment choices increases uptake and continuation.^59–61^ Educational tools could be used to inform PWH of their treatment options,^24,62^ empower them to choose what will work best for them, and facilitate equitable access.^13^ Materials should be developed with balanced information about all regimen options, since perceived coercion or loss of control may lower acceptability of treatment, regardless of treatment efficacy or effectiveness.^23,63^ Providers may also benefit from education around this newer biomedical technology.

In spite of limited prior knowledge of LAI ART, our findings suggest that there is interest in LAI ART among a subset of RWPA MCM clients in NYC and the Tri-County area. Based on programmatic data captured in eSHARE, 18 survey participants were on an LAI ART regimen at their most recent MCM visit as of October 2025; 16 of those were members of Group 2, which preferred LAI ART over daily pills. Participants were also more likely than other MCM clients to be on an LAI ART regimen at their most recent visit. As of the time of this writing, Cabenuva is only approved for PWH who have already achieved VS on their daily pill ART regimen, severely limiting the potential of LAI ART to bridge adherence gaps. One study that offered LAI ART to non-adherent ART clients found 80% achieved VS on LAI ART,^57^ indicating the potential of LAI ART to address VS failure on a daily pill ART regimen. The eligibility criterion of VS prior to LAI initiation may change with newer trial results on LAI ART efficacy specifically among those not virally suppressed under daily pill ART.^64^

Importantly, this study was conducted in a real-world MCM environment, with diversity across gender, racial and ethnic groups. One-quarter of participants self-reported imperfect adherence to daily pill ART and only three-quarters were virally suppressed. In contrast, in the Phase II and III trials for Cabenuva, participants had to be virally suppressed and were predominantly male and White.^65–67^ Assessing preferences among groups under-represented in clinical trials is essential to effective and equitable real-world implementation of LAI ART, and this study contributes to a growing body of work in this space.^17,45,53,54,68^

We note the following limitations. First, stated preferences from a DCE are based on hypothetical scenarios and may not align with future enacted choices. In addition, preferences are dynamic and are influenced by contextual factors;^69^ treatment option preferences vary between and within individuals, over time, and when new options become the established norm.^70^ However, previous research comparing stated preferences to revealed preferences suggests that preferences identified from DCEs do predict choices made in the real world,^71,72^ and our own data indicate that people who preferred LAI ART in the DCE were more likely to subsequently initiate LAI ART. Second, measurement error is common and may have arisen in our study if participants did not understand a question, if their responses were motivated by social desirability bias, or if they responded to questions without considering their true preferences. However, in our sensitivity analyses, excluding poor-quality responses made little difference in our findings. Third, we may have been underpowered when comparing some participant characteristics across LCA groups, as some cell counts are small. Finally, our study participants were a subset of clients in NY RWPA MCM programs who differed from non-participating peers in several characteristics. Their preferences may not be generalizable to the preferences of other PWH in NYC and the Tri-County area, in other jurisdictions, or in other care settings.

## Conclusions

Our study found that adult New York RWPA MCM clients had diverging preferences for ART regimens and limited familiarity with LAI ART. To maximize the impact of LAI ART among diverse populations in real-world settings, client and provider input should be used to develop easy-to-use educational and decision-making tools that empower clients to make informed ART regimen decisions. To facilitate regimen selection, we launched a pilot test of informational materials and a patient-provider decision-making tool in 2023.^35^ More work may be needed to adapt care systems to ensure access to and maintenance of newer treatment options for those who prefer them.^23,73,74^

## Data availability statement

De-identified discrete choice experiment data only will be made available upon reasonable request.

## Funding statement

Funding for the study was provided under NIMH grant R34MH126809. Funding for broader work and effort was provided under Health Resources and Services Administration (HRSA), grant number H89HA00015.

## Conflict of interest disclosure

Sarah Kulkarni, Denis Nash, and Rebecca Zimba have received funding from Pfizer for a project unrelated to the current project. No other authors have conflicts to declare.

## Ethics approval statement

The study methods were reviewed and approved by the Institutional Review Board of the New York City Department of Health and Mental Hygiene, protocol 20-096.

## Patient consent statement

Clients were required to review a consent form and provide electronic consent before they could begin the survey.

## Permission to reproduce material from other sources

N/A

## Author contributions

RZ, EAK, JC, MI, and DN contributed to the conception and design of the study; RZ, SK, TA, CE, MP, and MI contributed to the acquisition of data; RZ and TA analyzed the data; and all authors made substantial contributions to the interpretation of data. RZ, EAK, JC, MP, and MI drafted the manuscript; all authors were involved in revising it critically for important intellectual content. All authors approve of the version to be published and agree to be accountable for all aspects of the work.

## Acknowledgements

We gratefully acknowledge the contributions of Aimee Campbell, Chunki Fong, Gina Gambone, Honoria Guarino, Grace Herndon, Sarah Kozlowski, Javier Lopez-Rios, Monique Millington, Tyeirra Seabrook, the Assessing Perceptions and Preferences Around Long-acting Injectables (APPLI) study advisory board, and our Ryan White Part A medical case management partner sites and their clients. The study benefitted from the support of the HIV Health and Human Services Planning Council of New York, the New York City Health Department HIV Care and Treatment Program leadership and quality management staff and the Einstein-Rockefeller-CUNY Center for AIDS Research.

## Notes

### Author Declarations

The Institutional Review Board of the New York City Department of Health and Mental Hygiene gave ethical approval for this work under protocol 20-096.

