## Supplemental materials for "HIV Treatment and Program Preferences Among Ryan White Clients in New York City in the Era of Long-Acting Injectable ART: A Discrete Choice Experiment"

### Methods

Supplemental Table 1. Focus group participant demographic characteristics

| **Characteristic** | **Clients (N=5)**  **n (%)** | **Providers (N=16)**  **n (%)** |
| --- | --- | --- |
| **Age group** |  |  |
| 20-29 | 1 (20%) | 2 (12%) |
| 30-39 | 1 (20%) | 7 (44%) |
| 40-49 | 1 (20%) | 3 (19%) |
| 50-59 | 1 (20%) | 0 (0%) |
| 60+ | 1 (20%) | 4 (25%) |
| **Gender** |  |  |
| Man | 0 (0%) | 3 (19%) |
| Woman | 5 (100%) | 12 (75%) |
| Other | 0 (0%) | 1 (6%) |
| **Race/Ethnicity** |  |  |
| Black | 5 (100%) | 2 (12%) |
| Latino/a | 0 (0%) | 7 (44%) |
| Asian | 0 (0%) | 2 (13%) |
| White | 0 (0%) | 4 (25%) |
| Mixed race | 0 (0%) | 1 (6%) |
| **Agency location** |  |  |
| New York City | 4 (80%) | 12 (75%) |
| Tri-County | 1 (20%) | 4 (25%) |

*Sample size calculations*

To estimate the sample size for our DCE we used “n ≥ (500c)/(ta)”, where *n* is the number of participants, *c* is the maximum number of levels among all of the attributes, *t* is the number of choice tasks and *a* is the number of options per task.(Orme 2014; Mele 2008) This formula stipulates that each main-effect level appears at least 500 times within the survey design. The minimum sample size for the client DCE, given a maximum of four levels among our attributes, 12 choice tasks and 3 alternatives per task (including the None option) was 56. However, with only 56 participants, and assuming 33% of participants selected the None option, the estimated standard errors for the alternative-specific effects ranged from 0.10 to 0.14. Sawtooth recommends a design that achieves standard errors of 0.10 or smaller for alternative-specific effects. We set our target sample size to 200; again pessimistically assuming 33% of participants selected the None option, this yielded estimated standard errors for the alternative-specific effects ranging from 0.05 to 0.07.

Supplemental Table 2. Attributes and levels in the Client DCE for the APPLI Study

| **Attribute** | **Levels** | **Helper image** |
| --- | --- | --- |
| Type of ART Medication | An ART injection in each buttock **once a month** | 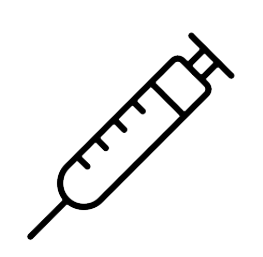 |
|  | An ART injection in each buttock **once every two months** | 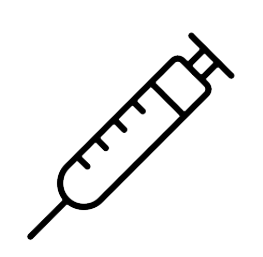 |
|  | Daily oral ART pill(s) | 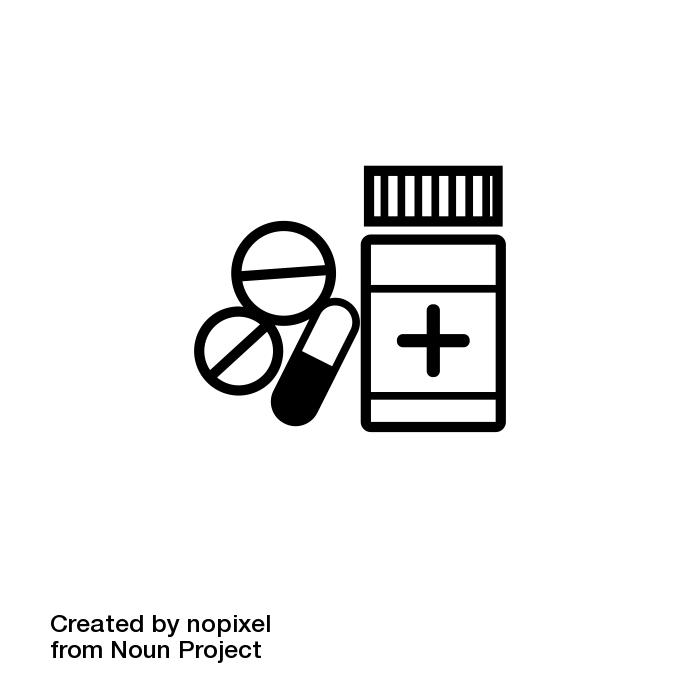 |
| Service location and mode | [Injection] **Injections** **by** **appointment** at clinic | 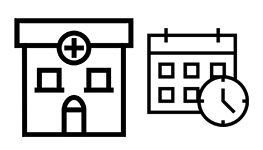 |
|  | [Injection] Walk-in for **injections** at clinic with **no appointment required** | 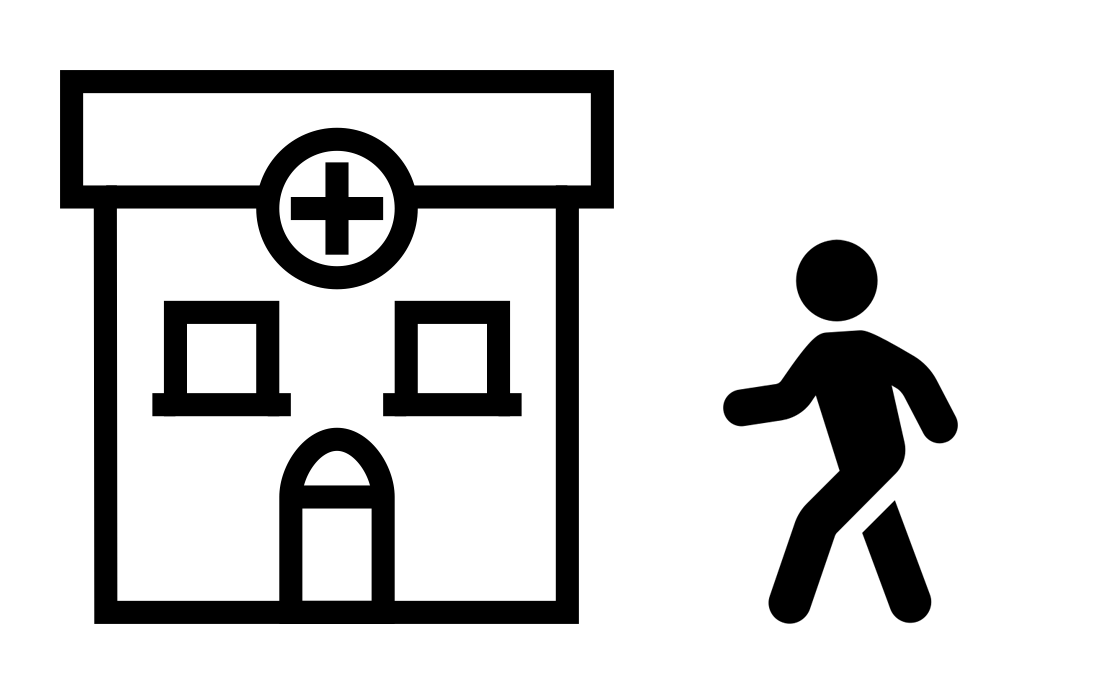 |
|  | [Injection] **Injections** **by appointment** at home | 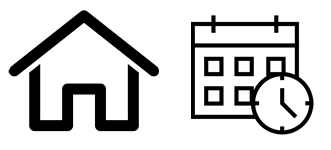 |
|  | [Pills] **ART pills** delivered to home | 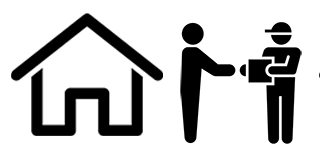 |
|  | [Pills] **ART** **pills** pickup from a pharmacy or other non-program location | 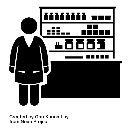 |
|  | [Pills] **ART** **pills** pickup from program location | 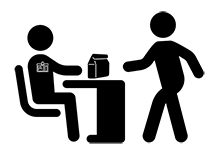 |
| Support | [Injection] Calls and texts to remind about **injection appointments** | 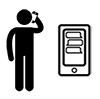 |
|  | [Injection] Free transportation provided to get **injections** | 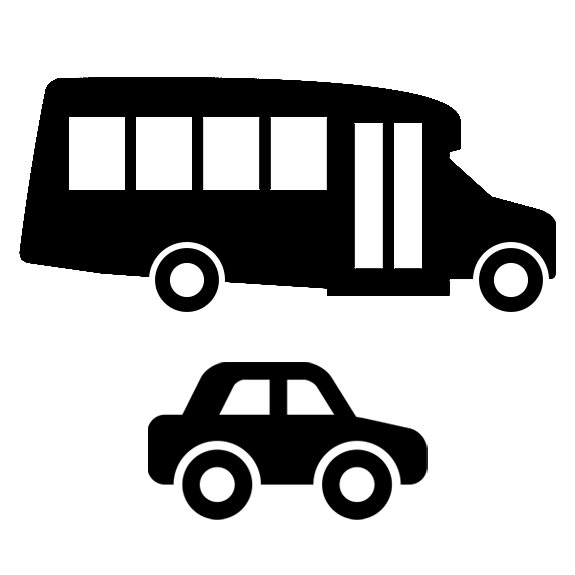 |
|  | [Injection] **Post-injection** follow-up calls from program staff | 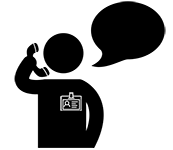 |
|  | [Injection] Peer support group for clients on or considering **long-acting injectable ART** | 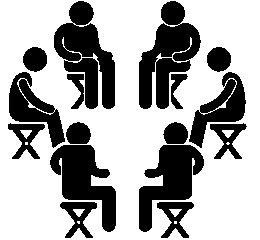 |
|  | [Pills] Calls and texts to remind about taking **ART pills** | 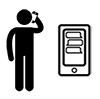 |
|  | [Pills] Free transportation provided to **primary care appointments** | 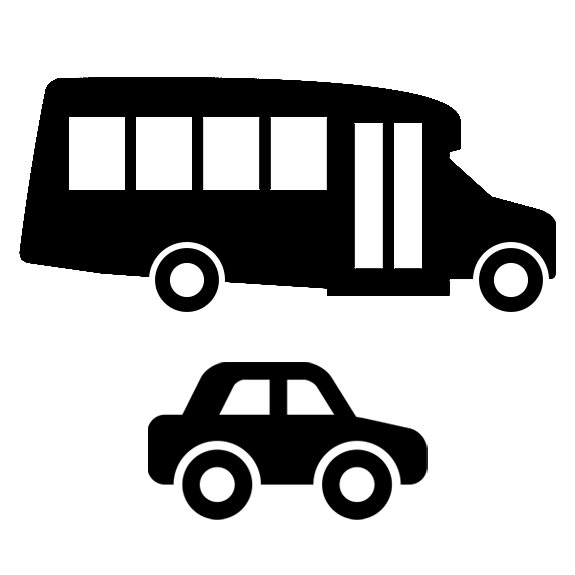 |
|  | [Pills] Peer support group to help with **ART pill adherence** | 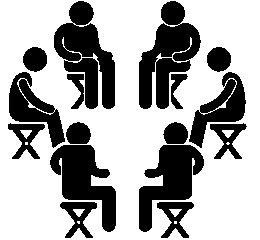 |
| Rewards | [Injection] **$10** gift card for every **injection visit** in the right time frame | 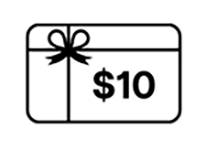 |
|  | [Injection] **$20** gift card for **every injection visit** in the right time frame | 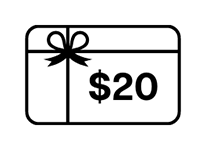 |
|  | [Pills] **$35** gift card every **6 months** for having **undetectable or suppressed viral load** | 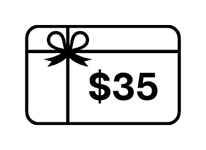 |
|  | [Pills] **$70** gift card every **6 months** for having **undetectable or suppressed viral load** | 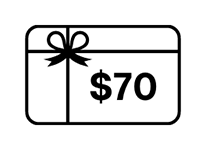 |
|  | [All] Rewards ceremony with program staff to celebrate meeting treatment goals | 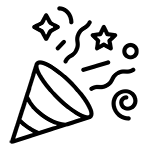 |

Supplemental Figure 1. Sample task from the Client DCE for the APPLI Study

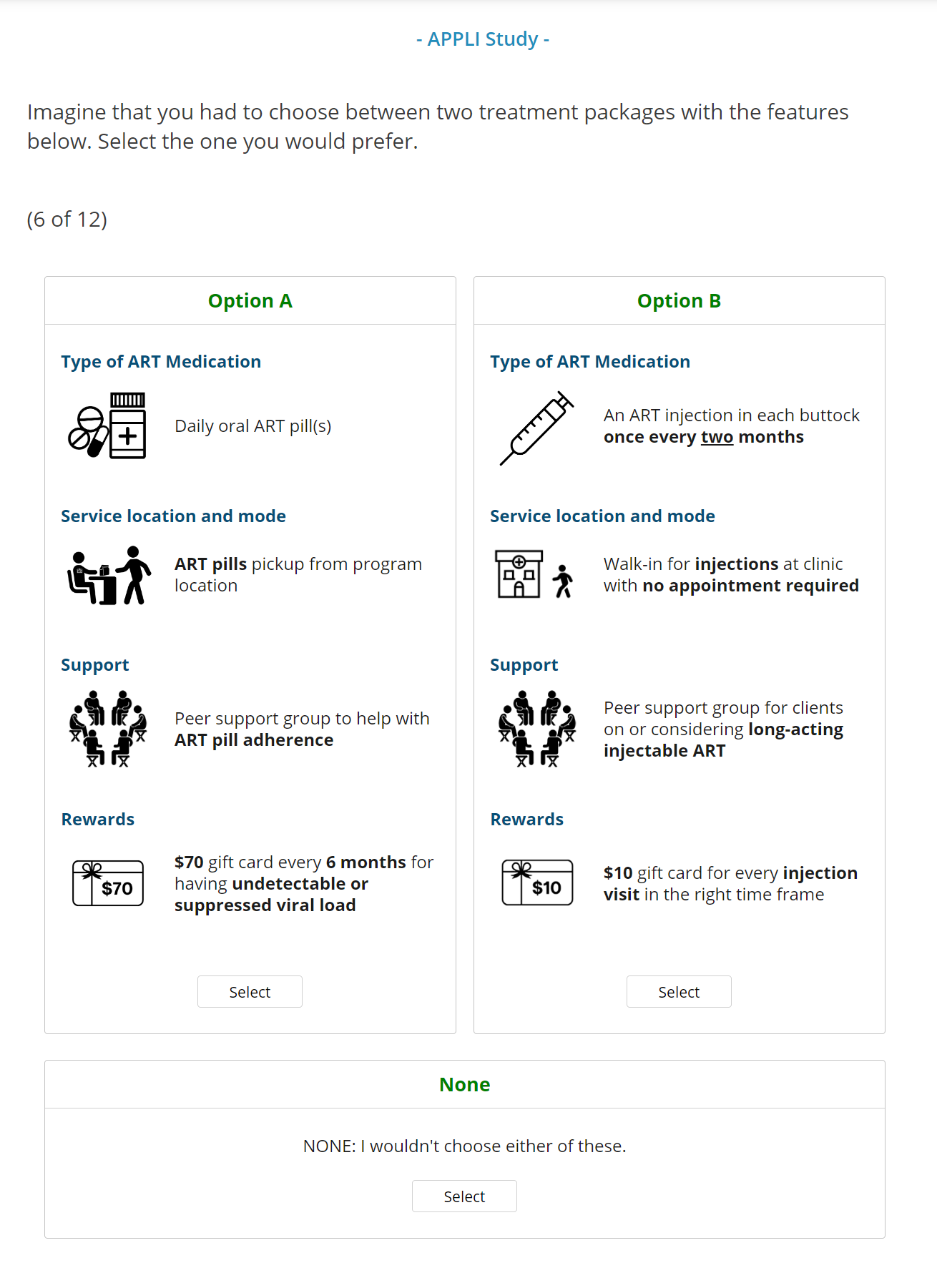

### Results

Supplemental Figure 2. Client eligibility and survey completion status for the APPLI Study

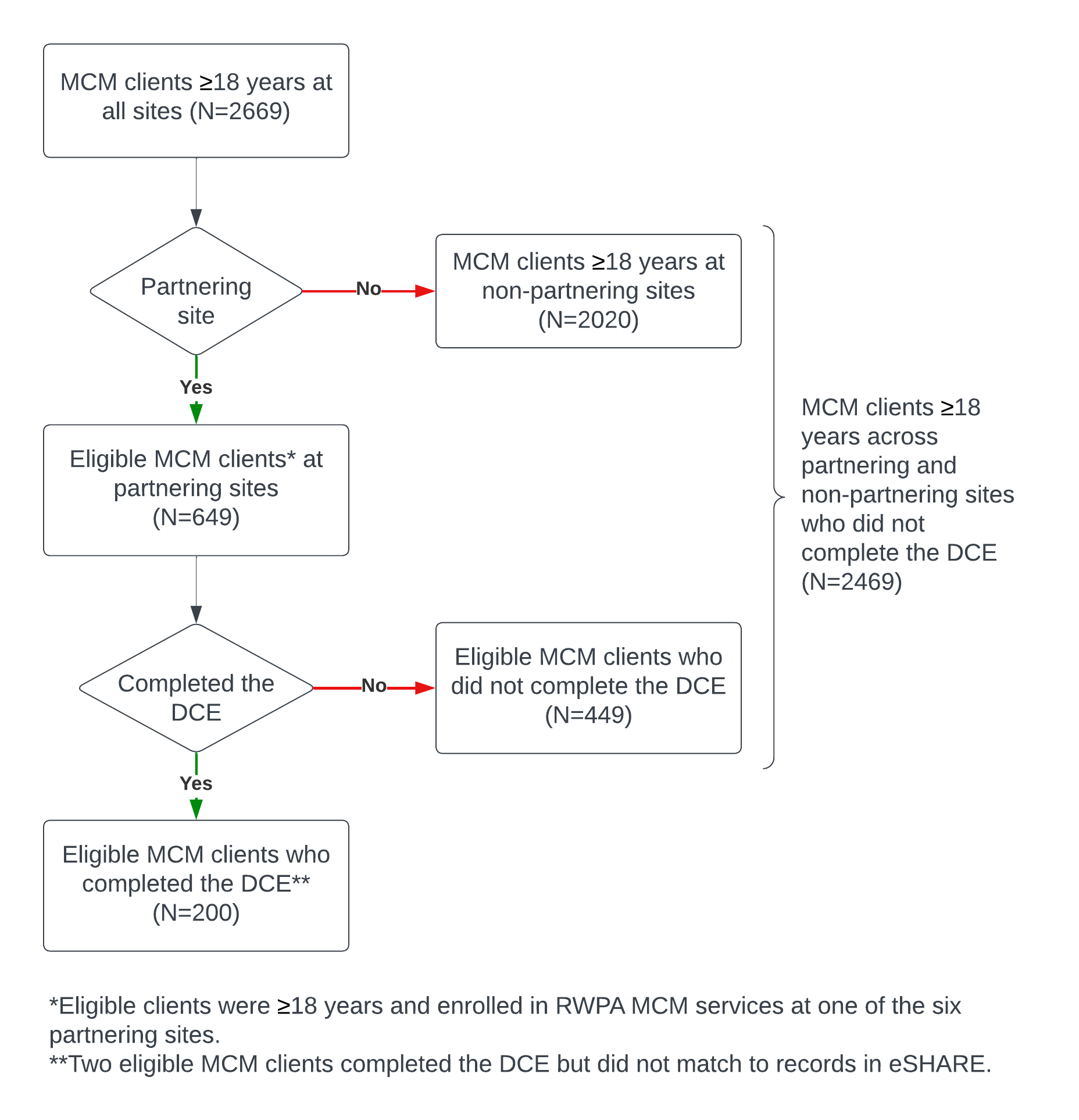

Supplemental Table 3. Comparison of characteristics between clients who completed the APPLI DCE and other clients at partnering MCM sites

|  | **Overall***^†^* N = 649 | **DCE completion status** | | |
| --- | --- | --- | --- | --- |
|  |  | **Completed DCE***^†,‡^* n=200 (31%) | **All other MCM clients***^†^* n=449 (69%) | **p-value** |
| **DEMOGRAPHIC VARIABLES** | | | | |
| **Age in years** | 54 (39, 62) | 55 (41, 62) | 54 (38, 62) | 0.7*^§^* |
| Missing/Unknown | 2 | 2 | 0 |  |
| **Age group in years** |  |  |  | 0.7*^¶^* |
| 18-29 | 53 (8.2%) | 17 (8.6%) | 36 (8.0%) |  |
| 30-39 | 115 (18%) | 30 (15%) | 85 (19%) |  |
| 40-49 | 101 (16%) | 33 (17%) | 68 (15%) |  |
| 50-59 | 162 (25%) | 47 (24%) | 115 (26%) |  |
| 60+ | 216 (33%) | 71 (36%) | 145 (32%) |  |
| Missing/Unknown | 2 | 2 | 0 |  |
| **Self-reported gender** |  |  |  | 0.4*^††^* |
| Cisgender man | 379 (59%) | 117 (59%) | 262 (58%) |  |
| Cisgender woman | 253 (39%) | 79 (40%) | 174 (39%) |  |
| Transgender, gender-non-conforming, or non-binary | 15 (2.3%) | 2 (1.0%) | 13 (2.9%) |  |
| Missing/Unknown | 2 | 2 | 0 |  |
| **Sexual identity** |  |  |  | 0.3*^¶^* |
| Homosexual, bisexual, pansexual, or queer | 182 (28%) | 50 (25%) | 132 (30%) |  |
| Straight/heterosexual | 461 (72%) | 147 (75%) | 314 (70%) |  |
| Missing/Unknown | 6 | 3 | 3 |  |
| **Race/Ethnicity***^‡‡^* |  |  |  | 0.5*^††^* |
| Latino/a | 257 (40%) | 87 (44%) | 170 (38%) |  |
| Asian/Pacific Islander | 3 (0.5%) | 0 (0%) | 3 (0.7%) |  |
| Black | 351 (54%) | 99 (50%) | 252 (56%) |  |
| White | 28 (4.3%) | 9 (4.5%) | 19 (4.2%) |  |
| Other/Multiracial | 8 (1.2%) | 3 (1.5%) | 5 (1.1%) |  |
| Missing/Unknown | 2 | 2 | 0 |  |
| **Primary language** |  |  |  | 0.3*^¶^* |
| English | 393 (61%) | 116 (59%) | 277 (62%) |  |
| Spanish | 191 (30%) | 67 (34%) | 124 (28%) |  |
| Haitian Creole | 41 (6.3%) | 9 (4.5%) | 32 (7.1%) |  |
| Other primary language | 22 (3.4%) | 6 (3.0%) | 16 (3.6%) |  |
| Missing/Unknown | 2 | 2 | 0 |  |
| **Highest level of schooling completed** |  |  |  | 0.085*^¶^* |
| High school/GED or lower | 476 (75%) | 137 (71%) | 339 (77%) |  |
| Greater than high school/GED | 158 (25%) | 57 (29%) | 101 (23%) |  |
| Missing/Unknown | 15 | 6 | 9 |  |
| **Employment status** |  |  |  | 0.4*^¶^* |
| Paid Employment | 220 (34%) | 75 (38%) | 145 (32%) |  |
| No Paid Employment | 293 (45%) | 86 (44%) | 207 (46%) |  |
| Out of Workforce | 131 (20%) | 36 (18%) | 95 (21%) |  |
| Missing/Unknown | 5 | 3 | 2 |  |
| **Housing status***^§§^* |  |  |  | >0.9*^¶^* |
| Stably Housed | 553 (85%) | 169 (85%) | 384 (86%) |  |
| Unstably Housed or Homeless | 94 (15%) | 29 (15%) | 65 (14%) |  |
| Missing/Unknown | 2 | 2 | 0 |  |
| **MCM ENROLLMENT AND AGENCY VARIABLES** | | | | |
| **Time since MCM enrollment in months** | 31.2 (14.5, 49.2) | 31.2 (11.8, 45.8) | 31.2 (14.8, 50.7) | 0.14*^§^* |
| Missing/Unknown | 2 | 2 | 0 |  |
| **Agency category** |  |  |  | <0.001*^††^* |
| Community health center | 279 (43%) | 107 (54%) | 172 (38%) |  |
| Community-based organization | 144 (22%) | 59 (30%) | 85 (19%) |  |
| Private hospital | 0 (0%) | 0 (0%) | 0 (0%) |  |
| Public hospital | 226 (35%) | 34 (17%) | 192 (43%) |  |
| **Agency location** |  |  |  | <0.001*^††^* |
| Bronx | 87 (13%) | 37 (19%) | 50 (11%) |  |
| Brooklyn | 226 (35%) | 34 (17%) | 192 (43%) |  |
| Manhattan | 77 (12%) | 25 (13%) | 52 (12%) |  |
| Queens | 0 (0%) | 0 (0%) | 0 (0%) |  |
| Staten Island | 0 (0%) | 0 (0%) | 0 (0%) |  |
| Tri-County | 259 (40%) | 104 (52%) | 155 (35%) |  |
| **MENTAL HEALTH AND SUBSTANCE USE VARIABLES** | | | | |
| **Psychiatric diagnoses (lifetime history, not mutually exclusive)***^¶¶^* |  |  |  |  |
| Depression or anxiety disorder | 279 (43%) | 96 (48%) | 183 (41%) | 0.067*^¶^* |
| Psychosis or bipolar disorder | 94 (15%) | 30 (15%) | 64 (14%) | 0.8*^¶^* |
| Post-traumatic stress disorder | 55 (8.5%) | 20 (10%) | 35 (7.8%) | 0.3*^¶^* |
| Other psychiatric diagnosis | 13 (2.0%) | 5 (2.5%) | 8 (1.8%) | 0.6*^††^* |
| Missing/Unknown (for all psychiatric diagnoses) | 2 | 2 | 0 |  |
| **Recent problem substance use***^†††^* | 99 (16%) | 18 (10%) | 81 (19%) | 0.007*^¶^* |
| Missing/Unknown | 40 | 21 | 19 |  |
| **HIV CLINICAL AND TREATMENT VARIABLES** | | | | |
| **Time since HIV diagnosis in years** | 17.0 (7.8, 27.1) | 18.2 (8.8, 27.5) | 16.5 (7.2, 26.8) | 0.14*^§^* |
| Missing/Unknown | 36 | 15 | 21 |  |
| **ART regimen proximal to date of survey** |  |  |  | 0.043*^††^* |
| Daily oral ART | 621 (96%) | 196 (99%) | 425 (95%) |  |
| LAI ART | 4 (0.6%) | 0 (0%) | 4 (0.9%) |  |
| Other, unspecified ART | 11 (1.7%) | 2 (1.0%) | 9 (2.0%) |  |
| Not on ART | 11 (1.7%) | 0 (0%) | 11 (2.4%) |  |
| Missing/Unknown | 2 | 2 | 0 |  |
| **Percent adherence***^‡‡‡^* |  |  |  | 0.10*^¶^* |
| Below 50% | 41 (6.6%) | 7 (3.6%) | 34 (8.0%) |  |
| 50-69% | 31 (5.0%) | 6 (3.1%) | 25 (5.9%) |  |
| 70-89% | 35 (5.6%) | 9 (4.6%) | 26 (6.1%) |  |
| 90-94% | 38 (6.1%) | 13 (6.6%) | 25 (5.9%) |  |
| 95-99% | 39 (6.3%) | 10 (5.1%) | 29 (6.8%) |  |
| 100% | 437 (70%) | 151 (77%) | 286 (67%) |  |
| **Receipt of directly observed therapy within MCM***^§§§^* | 130 (20%) | 50 (25%) | 80 (18%) | 0.035*^¶^* |
| **Most recent ART regimen***^¶¶¶^* |  |  |  | 0.12*^††^* |
| Daily oral ART | 586 (91%) | 175 (88%) | 411 (92%) |  |
| LAI ART | 39 (6.0%) | 18 (9.1%) | 21 (4.7%) |  |
| Other, unspecified ART | 8 (1.2%) | 1 (0.5%) | 7 (1.6%) |  |
| Not on ART | 14 (2.2%) | 4 (2.0%) | 10 (2.2%) |  |
| Missing/Unknown | 2 | 2 | 0 |  |
| **Viral load (copies/mL)***^††††^* |  |  |  | 0.009*^¶^* |
| <200 | 519 (80%) | 171 (86%) | 348 (78%) |  |
| >=200 | 128 (20%) | 27 (14%) | 101 (22%) |  |
| Missing/Unknown | 2 | 2 | 0 |  |
| **CD4 category (cells/mm3)***^††††^* |  |  |  | 0.049*^¶^* |
| <200 | 197 (30%) | 73 (37%) | 124 (28%) |  |
| 200-349 | 85 (13%) | 23 (12%) | 62 (14%) |  |
| 350-499 | 100 (15%) | 22 (11%) | 78 (17%) |  |
| 500+ | 265 (41%) | 80 (40%) | 185 (41%) |  |
| Missing/Unknown | 2 | 2 | 0 |  |
| **Resistance testing history** |  |  |  | <0.001*^¶^* |
| Resistance testing in past year | 61 (9.4%) | 16 (8.1%) | 45 (10%) |  |
| Resistance testing >1 year ago | 253 (39%) | 58 (29%) | 195 (43%) |  |
| No resistance testing reported | 333 (51%) | 124 (63%) | 209 (47%) |  |
| Missing/Unknown | 2 | 2 | 0 |  |
| **Time since last HIV ART resistance test in months***^‡‡‡‡^* | 42.0 (16.6, 79.0) | 54.3 (15.2, 120.8) | 39.7 (16.8, 74.9) | 0.2*^§^* |
| Abbreviation: ART = antiretroviral therapy; DCE = discrete choice experiment; GED = general equivalency diploma; MCM = medical case management. | | | | |
| *^†^* Median (Q1, Q3); n (%) | | | | |
| *^‡^* DCE and programmatic records could not be linked for two clients who completed the DCE. | | | | |
| *^§^* Wilcoxon rank sum test | | | | |
| *^¶^* Pearson’s Chi-squared test | | | | |
| *^††^* Fisher’s exact test | | | | |
| *^‡‡^* Black, White, Asian/Pacific Islander, and Other/Multiracial race categories exclude Latino/a ethnicity. People with the ethnicity Latino/a are grouped in the Latino/a category regardless of their race classification | | | | |
| *^§§^* ‘Stable’ defined as any permanent housing situation. ‘Unstable’ defined as any temporary/transitional housing situation or being homeless. Reported as of client’s latest available eSHARE assessment before date of survey completion OR January 31, 2023. | | | | |
| *^¶¶^* Any report of a psychiatric diagnosis on any eSHARE assessment before the date of survey completion OR January 31, 2023. | | | | |
| *^†††^* Recent report of binge drinking, heroin, meth, cocaine, or recreational prescription drug use in last 3 months. Reported as of client’s latest available eSHARE assessment before date of survey completion OR January 31, 2023. | | | | |
| *^‡‡‡^* Self-reported percent of the past 30 days on which client took their ART pills, as of client’s latest available eSHARE assessment before date of survey completion OR January 31, 2023. | | | | |
| *^§§§^* Includes directly observed therapy received from MCM agencies in the last decade. | | | | |
| *^¶¶¶^* As of October 28, 2025. | | | | |
| *^††††^* Most recent lab values or self-reported values recorded in eSHARE before date of survey completion OR January 31, 2023. | | | | |
| *^‡‡‡‡^* Among clients who had evidence of resistance testing. | | | | |

Supplemental Table 4. Model fit statistics and latent class sizes

| **DCE** | **Groups** | **Group size range** | **Log-likelihood** | **Percent Certainty^†^** | **AIC** | **BIC** |
| --- | --- | --- | --- | --- | --- | --- |
| Original analysis | Null | 0 | -2636.67 |  |  |  |
|  | 1 | 200 | -2383.87 | 9.59 | 4799.74 | 4892.27 |
|  | 2 | 86–114 | -1897.70 | 28.03 | 3861.40 | 4052.25 |
|  | 3 | 16–108 | -1715.98 | 34.92 | 3531.96 | 3821.12 |
|  | 4 | 16–68 | -1617.18 | 38.67 | 3368.37 | 3755.84 |
|  | 5 | 15–75 | -1570.14 | 40.45 | 3308.28 | 3794.07 |
| Sensitivity analysis excluding poor quality responses^‡^ | Null | 0 | -2122.52 |  |  |  |
|  | 1 | 161 | -1838.94 | 13.36 | 3709.87 | 3798.93 |
|  | 2 | 75–86 | -1371.37 | 35.39 | 2808.75 | 2992.44 |
|  | 3 | 52–55 | -1259.73 | 40.65 | 2619.45 | 2897.77 |
|  | 4 | 8–52 | -1183.94 | 44.22 | 2501.87 | 2874.82 |
|  | 5 | 9–46 | -1156.00 | 45.54 | 2480.00 | 2947.57 |
| **^†^** Percent certainty between 0.2 and 0.4 indicates a good model fit (Hauber et al. 2016).  ^‡^ Poor quality responses included those that exhibited straightlining, were completed faster than the fastest 5^th^ percentile of responses, or had lower root likelihood values than the 95^th^ percentile of random responses. | | | | | | |

*Respondent quality and sensitivity analysis*

To find an appropriate RLH cutoff value for identifying potentially inconsistent responses, we analyzed 200 randomly generated responses and found the 95^th^ percentile RLH value, 0.558. A total of 23 (11%) responses fell below this threshold. We identified nine responses with straightlining: one client selected all option A, one client selected all option B, and seven clients selected the none option for all choice tasks. The average time to complete the twelve choice tasks was seven minutes (range 1.2 minutes to 28 minutes); nine clients’ completion times were faster than the fastest 5^th^ percentile completion time, 1.9 minutes. We re-ran the latent class analysis for the subset of clients with any of the above respondent quality indicators, excluding 39 participants. Apart from the smaller sample size, the overall composition of the two latent classes retained similarity across analyses; seven observations moved from Group 2 in the primary analysis to Group 1 in the sensitivity analysis.

The overall preference for daily pill ART in Group 1 (n=75, 47%) and LAI ART in Group 2 (n=86, 53%) persisted in the sensitivity analysis. Though the numerical values of the utilities changed, the levels’ relative ranking was mostly conserved within each attribute (see Supplemental Table 4 and Supplemental Figure 3). There were two differences in the order of levels between the original and sensitivity analyses. In Group 1, in the original analysis, the calls and texts to remind about taking ART pills level in the Support attribute was ranked higher than the peer support group level, though they had very similar values (-8.8 versus -9.1 utiles). In contrast, in the sensitivity analysis, the calls and texts to remind about taking ART pills level was ranked lower than the peer support group level (-14.4 versus 4.3 utiles). Similarly, in Group 2, the injections by appointment at home level in the Service Location and Mode attribute was ranked higher than the injections by appointment at clinic level (5.9 versus 3.6 utiles), whereas in the sensitivity analysis injections by appointment at home level was ranked lower than the injections by appointment at clinic level (3.0 versus 5.8 utiles). All other levels retained the same order of preference as in the primary analysis, though there were some differences in magnitude.

Supplemental Table 5. Sensitivity analysis^†^ of preferences of clients from the APPLI DCE estimated with alternative-specific utilities from latent class multinomial logit regression analysis, two-group solution, overall and by latent class group

| **Attribute** | **Levels** | **Overall** N = 161 Mean (95% CI) | **Latent class analysis groups^‡^** | |
| --- | --- | --- | --- | --- |
|  |  |  | **Group 1, tend to prefer pills** n = 75 (47%) Mean (95% CI) | **Group 2, tend to prefer LAI** n = 86 (53%) Mean (95% CI) |
| Type of ART Medication | Daily oral ART pill(s) | 48.8 (21.9, 75.8) | 233.0 (230.4, 235.7) | -111.8 (-115.3, -108.4) |
|  | An ART injection in each buttock once every two months | -2.1 (-16.0, 11.9) | -97.5 (-99.0, -95.9) | 81.1 (79.7, 82.5) |
|  | An ART injection in each buttock once a month | -46.8 (-59.8, -33.8) | -135.6 (-136.7, -134.5) | 30.7 (28.7, 32.7) |
| Service Location and Mode – Pills | ART pills delivered to home | 36.9 (36.6, 37.2) | 37.0 (36.7, 37.3) | 36.7 (36.2, 37.3) |
|  | ART pills pickup from program location | -20.7 (-21.9, -19.5) | -28.6 (-28.6, -28.6) | -13.8 (-14.3, -13.4) |
|  | ART pills pickup at pharmacy or other non-program location | -16.2 (-17.3, -15.0) | -8.4 (-8.7, -8.2) | -22.9 (-23.0, -22.8) |
| Support – Pills | Free transportation provided to primary care appointments | 13.2 (12.8, 13.7) | 10.1 (10.0, 10.3) | 16.0 (15.8, 16.1) |
|  | Peer support group to help with ART pill adherence | -8.1 (-9.9, -6.3) | 4.3 (4.0, 4.6) | -18.9 (-19.0, -18.9) |
|  | Calls and texts to remind about taking ART pills | -5.1 (-6.5, -3.8) | -14.4 (-14.5, -14.3) | 3.0 (2.8, 3.2) |
| Rewards – Pills | $70 gift card every 6 months for having undetectable or suppressed viral load | 62.2 (60.4, 64.0) | 50.3 (49.6, 50.9) | 72.5 (71.8, 73.3) |
|  | $35 gift card every 6 months for having undetectable or suppressed viral load | -1.1 (-2.6, 0.4) | -11.4 (-11.5, -11.2) | 7.9 (7.7, 8.1) |
|  | Rewards ceremony with program staff to celebrate meeting treatment goals | -61.1 (-64.4, -57.8) | -38.9 (-39.7, -38.1) | -80.4 (-81.1, -79.8) |
| Service Location and Mode – Injections | Injections by appointment at clinic | 7.5 (7.2, 7.8) | 9.5 (9.4, 9.5) | 5.8 (5.7, 6.0) |
|  | Walk-in for injections at clinic with no appointment required | -5.2 (-5.8, -4.6) | -1.1 (-1.2, -1.0) | -8.8 (-8.9, -8.8) |
|  | Injections by appointment at home | -2.3 (-3.2, -1.4) | -8.4 (-8.5, -8.3) | 3.0 (2.9, 3.1) |
| Support – Injections | Free transportation provided to get injections | 41.4 (36.1, 46.7) | 5.1 (4.2, 6.0) | 73.1 (72.9, 73.2) |
|  | Post-injection follow-up calls from program staff | -8.1 (-11.2, -4.9) | 13.5 (13.1, 13.9) | -26.9 (-27.1, -26.7) |
|  | Calls and texts to remind about injection appointments | 2.9 (2.2, 3.6) | 7.7 (7.7, 7.8) | -1.3 (-1.4, -1.2) |
|  | Peer support group for clients on or considering long-acting injectable ART | -36.3 (-37.7, -34.8) | -26.4 (-26.8, -25.9) | -44.9 (-45.3, -44.5) |
| Rewards – Injections | $20 gift card for every injection visit in the right time frame | 56.1 (54.7, 57.6) | 46.7 (46.1, 47.3) | 64.4 (63.6, 65.1) |
|  | $10 gift card for every injection visit in the right time frame | -1.1 (-1.2, -0.9) | 0.3 (0.3, 0.3) | -2.2 (-2.2, -2.2) |
|  | Rewards ceremony with program staff to celebrate meeting treatment goals | -55.1 (-56.3, -53.8) | -47.0 (-47.5, -46.4) | -62.1 (-62.9, -61.4) |
| None | NONE | -258.1 (-298.7, -217.6) | 19.1 (12.6, 25.5) | -499.9 (-500.3, -499.5) |
| Abbreviations: ART = antiretroviral therapy, CI = confidence interval, LAI = long-acting antiretroviral  ^†^ Excluding poor quality responses included those that exhibited straightlining, were completed faster than the fastest 5th percentile of responses, or had lower root likelihood values than the 95th percentile of random responses.  ^‡^ All group comparisons statistically significant at p<0.001 using the Wilcoxon rank sum test. | | | | |

Supplemental Figure 3. Sensitivity analysis excluding poor quality responses^†^ compared with the original analysis: preferences of clients from the APPLI DCE estimated with alternative-specific utilities from latent class multinomial logit regression analysis, two-group solution

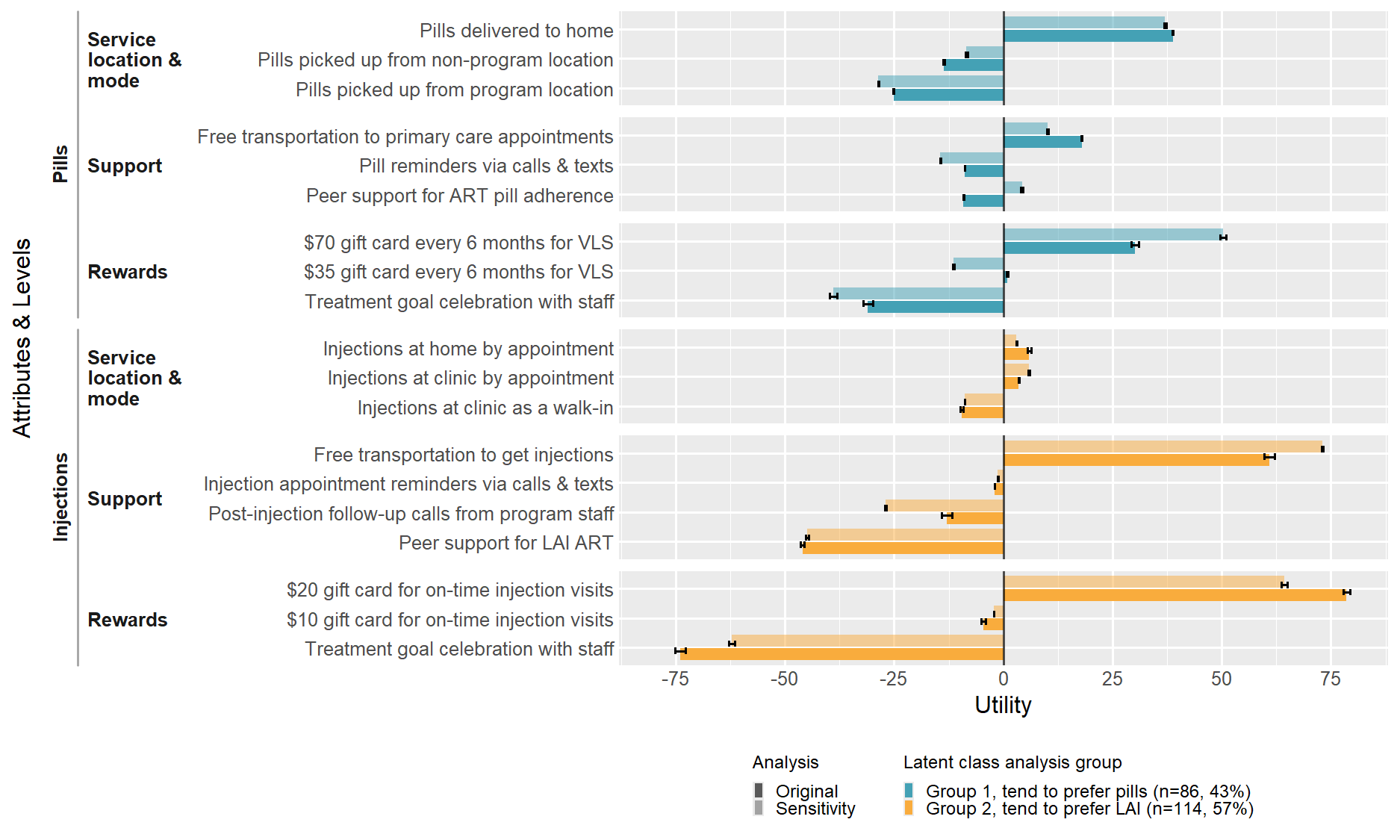

^†^ Poor quality responses included those that exhibited straightlining, were completed faster than the fastest 5^th^ percentile of responses, or had lower root likelihood values than the 95^th^ percentile of random responses.

Orme, Bryan K. 2014. *Getting Started with Conjoint Analysis*. Research Publishers, LLC.
